# Sirolimus in Patients with Overgrowth, with or without Vascular Malformations: The SIPA-SOS Study

**DOI:** 10.64898/2026.09.25.26364046

**Authors:** Gabriella Cericola, Beate Häberle, Julia Haehl, Sebastian Berg, Simon Leschka, Susanne Grüninger, Claudia Schmoor, Sebastian Wirtz, Annegret Holm, Charlotte M. Niemeyer, Alexander Puzik, Martin Zenker, Moritz Wildgruber, Jochen Rößler, Friedrich G. Kapp

## Abstract

**Background:** Localized or segmental overgrowth is a rare condition that may co-occur with vascular malformations. These diseases generally are caused by somatic mutations in the PI3K/AKT/mTOR pathway. Most lesions are progressive and may cause symptoms including pain, functional impairment, and disfigurement. Treatment options are limited, especially for the overgrowth component.

**Objective:** To evaluate the efficacy and safety of the mTOR inhibitor sirolimus in patients with overgrowth with or without a vascular malformation over a 6-months treatment period.

**Methods:** We conducted a single-arm, open-label, phase II study including 18 patients (12 pediatric, six adult) with overgrowth, all of whom initiated oral sirolimus. 16/18 (88.9%) participants received oral sirolimus for 6 months, with dosing adjusted to achieve target blood trough levels of 3–8 ng/mL. Primary outcome was radiological response at six months assessed by the Response Evaluation Criteria in Solid Tumors (RECIST) 1.1. Secondary outcome parameters were evaluated at three, six and nine months and included size and visual appearance of the overgrowth, quality of life (QoL), pain intensity, performance status, and safety.

**Results:** No patient achieved complete or partial radiological response. 17 of 18 patients (94.4%) maintained stable disease at month six; one was not evaluable. QoL measured with WHOQOL-BREF improved modestly in patients aged >= 18 years (n=5, mean difference to baseline +9.3 (95%-CI: -0.9 to 19.5) in physical and +9.2 (95%-CI: 0.7 to 17.7) in psychological domains at month 6). In pediatric participants, no relevant improvement in KINDL total scores was observed over the study period. No significant changes occurred in pain scores. Performance status measured by the Lansky/Karnofsky scales showed minor improvement (mean +3.5 points (95%-CI: 1.0 to 6.1)). All patients experienced at least one adverse event, but no serious adverse events or CTCAE v5.0 grade ≥3 toxicities occurred. Sirolimus was well tolerated. Six patients decided to resume sirolimus after study completion.

**Conclusions:** Sirolimus induced minor lesion shrinkage in a subset of patients, although this did not meet the predefined radiological RECIST endpoint. However, sirolimus can lead to functional improvement in some adults, which may indicate that the primary radiological outcome did not capture patient-perceived benefit.

## INTRODUCTION

Local or segmental overgrowth can lead to disfigurement, pain and functional impairment, which often begins in childhood and persists lifelong. Overgrowth can be accompanied or be caused by vascular malformations thus comprising a heterogeneous group of disorders [1]. Conditions with overgrowth with or without a vascular malformation include disease entities within the PIK3CA-related overgrowth spectrum (PROS), such as CLOVES syndrome (congenital lipomatous overgrowth, vascular malformations, epidermal nevi, and scoliosis/skeletal/spinal anomalies) and Klippel–Trénaunay syndrome, which show a segmental distribution. Other examples include entities like Proteus syndrome (due to a somatic mutation in *AKT1*) and *PTEN* hamartoma tumor syndrome. The latter share aberrant activation of the PI3K/AKT/mTOR pathway but differ not only in the underlying mutation but also in their phenotype [2].

In PROS, somatic activating variants in *PIK3CA* lead to constitutive PI3K signaling with increased downstream AKT and mTOR activity resulting in overgrowth of adipose, connective, and vascular tissues. Similarly, gain-of-function variants in *AKT1* and loss-of-function variants in *PTEN* converge on hyperactivation of the same pathway, providing a unifying mechanistic framework for a spectrum of vascular malformations and overgrowth. Clinically, patients frequently experience pain, recurrent infections or bleeding from malformations, and significant psychosocial burden related to disfigurement and functional limitations [3].

To date, pharmacological treatment options for overgrowth with or without vascular malformations remain limited. Surgical debulking and interventional procedures such as sclerotherapy are frequently required but are often associated with recurrence, morbidity, and incomplete symptom control. They predominantly target the malformation and, to a lesser degree the overgrowth. The advent of targeted therapies has changed the therapeutic landscape, yet no universally accepted and evidence-based systemic treatment exists for most overgrowth phenotypes. Sirolimus, an mTOR inhibitor, emerged as a promising option after early reports suggested clinical benefit in complicated vascular anomalies, particularly slow-flow malformations [4]. However, subsequent prospective studies in PROS and vascular malformations indicated that while sirolimus may be most effective in lymphatic malformations, it seems to primarily slow pathological tissue growth rather than induce major regression of existing overgrowth [3,5].

In contrast, the role of sirolimus with respect to radiological response in segmental overgrowth with or without vascular malformations remains incompletely defined. Given its favorable safety profile and well-established use in vascular anomalies across extended treatment durations, sirolimus remains an attractive candidate for further investigation in patients with extensive overgrowth. Whether mTOR inhibition can modify the course of overgrowth warrants evaluation in a prospective, standardized setting. [2,3,5].

Therefore, this multicenter, phase II trial aimed to evaluate the efficacy and safety of sirolimus in pediatric (≥3 years of age) and adult patients with overgrowth with and without vascular malformations. The primary endpoint was RECIST-based magnetic resonance imaging (MRI) assessment of lesion size, complemented by a comprehensive panel of patient-reported and functional outcomes as secondary endpoints.

## METHODS

This was a phase II, open⍰label, single⍰arm trial initiated at five tertiary centers in Germany, two of which recruited patients. Participants received sirolimus for six months followed by a three months off-treatment follow-up period to assess durability of treatment response.

### Participants

Eligible participants were children or adults (age ≥ 3 years) with overgrowth with or without a vascular malformation. This included patients with PROS (e.g. CLOVES syndrome or facial infiltrating lipomatosis), Proteus syndrome, PTEN hamartoma syndrome, or large vascular malformations associated with clinically significant and measurable volume increase of corresponding body parts. Each patient had at least one lesion ≥3 cm measurable on MRI. Study participation was offered to patients after discussion in an interdisciplinary board for vascular anomalies. All participants (or their legal guardians) provided written informed consent. The study was conducted in accordance with the guidelines of Good Clinical Practice and the Declaration of Helsinki and was approved by the local Ethics Committee (approval number 21-1037_1-AMG-ff multi; EudraCT 2015-005416-15; DRKS00010085). The post-study follow-up was performed as part of the registry for vascular malformations and tumors in Freiburg (approval number 21-1200).

### Treatment and Assessments

Patients received oral sirolimus at a starting dose of 1.6 mg/m^2^ per day. In patients under 16 years, the daily dose was divided into two doses of each 0.8 mg/m^2^ (morning and evening), while patients aged ≥16 years received the full dose once daily. Treatment continued for six months, with dose adjustments to maintain target trough sirolimus levels of 3–8 ng/mL. After the six months treatment period, patients were followed for an additional three months off therapy. Baseline evaluations included MRI of target lesions, standardized photographs for assessment of disfigurement – defined as lesion-related alteration in contour, bulk, or symmetry – quality-of-life (QoL) questionnaires, pain scales, and laboratory tests. These assessments were repeated at three, six, and nine months.

The primary efficacy endpoint was objective response at six months, defined as the achievement of a complete response (CR) or partial response (PR) of target lesions on MRI, according to RECIST 1.1 criteria [6]. RECIST 1.1 defines CR as disappearance of all target lesions, PR as ≥30% decrease in summed lesion diameter, progressive disease [PD] as ≥20% increase in lesion size or new lesions, and stable disease (SD) for changes in between. A central radiological review of MRI scans was performed to assess responses.

Secondary endpoints, assessed at months three, six and nine, included changes in lesion size (evaluated by centralized radiological MRI assessment) and appearance (evaluated by centralized independent assessment of photographs obtained during the study by three evaluators who were blinded to patient identifiers, local clinical assessments, clinical narrative data, and time point of photograph (except baseline)). Patient-reported QoL (using WHOQOL-BREF [7,8] for adults and KINDL [9] for children, along with Lansky/Karnofsky [9,10] performance status scales), pain intensity (10-point visual analogue scales appropriate to age), and neuropsychological status (Strengths and Difficulties Questionnaire, SDQ [11]). Serum biomarkers related to overgrowth, such as insulin-like growth factor 1 (IGF-1), IGF binding protein-3 (IGFBP-3) were measured at baseline and during treatment to explore pharmacodynamic effects. Safety assessments included regular monitoring of laboratory parameters and recording of all adverse events, graded by CTCAE v5.0. Drug compliance was monitored via patient diaries and periodic measurement of sirolimus trough levels.

### Statistical Analysis

The study employed a single-arm phase II design with a planned sample size of 18 patients. Sample size calculations were performed using the exact binomial distribution and were based on the following assumptions and requirements for the primary endpoint objective response (CR or PR): Under the assumption that the response probability in the absence of treatment would be near zero, the study should have a power of 90% to show at a one-sided significance level of 0.05 that the response probability under treatment with Sirolimus is larger than 2%. Under the assumption that this probability is 20% or higher in reality, the required sample size is 18 patients, and the treatment would be considered promising if at least two or more patients achieved a CR or PR at six months. All patients who received at least one dose of sirolimus were included in the full analysis set for efficacy and safety analyses. The probability of response was estimated as the relative frequency of patients who achieve a CR or PR in the full analysis set and was reported with two-sided 90% (corresponding to the planned test procedure) and with two-sided 95% confidence intervals (CI) based on the exact binomial distribution. Changes in continuous secondary outcomes from baseline were summarized with mean differences and 95% CIs. No formal hypothesis testing was performed on secondary endpoints due to the exploratory nature of the study.

## RESULTS

### Patient Population and Disposition

A total of 18 patients (12 females and six males) were enrolled between October 2021 and October 2022 and started sirolimus treatment. The median age at enrolment was 15 years (range 6–35). Six participants were adults (≥18 years) and 12 were children. **(Table 1)**. Baseline disease characteristics were heterogeneous; most patients had extensive overgrowth involving the limbs or craniofacial regions, often with prior surgical interventions, reflecting a severely affected population. Genetic testing was performed in 10 patients – either on lesional tissue or on blood (in case of a suspected germline variant) – and revealed pathogenic or likely pathogenic variants in *PIK3CA* in seven patients (all somatic), *PTEN* in two patients (one somatic and one germline), and in *TEK* in 1 patient (somatic) **(Table 2)**.

**Table 1.** Demographic Characteristics of the Study Population.

| Characteristic | n/N | % |
| --- | --- | --- |
| Total patients | 18/18 | 100 |
| Sex – Female | 12/18 | 66.7 |
| Sex – Male | 6/18 | 33.3 |
| Pediatric patients (<18 years) | 12/18 | 66.7 |
| Adult patients (≥18 years) | 6/18 | 33.3 |

Baseline demographic characteristics of patients enrolled in the SIPA-SOS study.

**Table 2.** Baseline Disease, Clinical Characteristics and Genetic Findings.

|  | n/N | % |
| --- | --- | --- |
| **Disease** |  |  |
| - Mixed capillary-lymphatic-venous malformation | 4/18 | 22.2 |
| - Infiltrating lipomatosis | 3/18 | 16.7 |
| - PTEN-associated vascular malformation | 2/18 | 11.1 |
| - Venous malformation | 4/18 | 22.2 |
| - Lymphatic malformation | 4/18 | 22.2 |
| - Fibroadipose vascular anomaly | 1/18 | 5.6 |
| **Clinical characteristics** |  |  |
| Progressive overgrowth | 10/18 | 55.6 |
| Proportional overgrowth | 8/18 | 44.4 |
| Disfigurement at baseline | 14/18 | 77.8 |
| Pain at baseline | 9/18 | 50.0 |
| Functional impairment | 8/18 | 44.4 |
| **Genetic findings** |  |  |
| Genetic testing performed | 10/18 | 55.5 |
| - PIK3CA | 7/10 | 70 |
| - PTEN | 2/10 | 20 |
| - TEK | 1/10 | 10 |

Clinical manifestations of overgrowth syndromes and results of genetic testing at baseline. Disfigurement at baseline was defined as a visible lesion-related alteration in body contour, symmetry, or external appearance documented on clinical examination and/or standardized photography. Functional impairment was defined as lesion-related limitation of organ function or activities of daily living, for example impaired mobility, breathing, feeding, speech, vision, or limb use, as documented clinically at baseline.

16 out of 18 patients (89%) completed the full six-months treatment course. Two patients discontinued sirolimus during the treatment phase: one (01-10) withdrew consent in the first month (after ∼3 weeks) and one (01-15) stopped at around 5 months due to recurrent aphthous ulcers and lack of perceived benefit. Two additional patients (01-12 und 01-13) withdrew from the study due to worsening symptoms within one month following the cessation of the study medication at month 6. One additional patient (01-01) switched to another clinical trial after completion of the protocol-defined follow-up assessments and was therefore considered fully documented. Consequently, 14 of 18 patients (78%) completed the entire nine-month study duration with final assessments.

Adherence to sirolimus therapy was high. The mean cumulative dose administered was approximately 233 mg/m^2^ per patient, corresponding to 86.7% of the planned total dose. Trough sirolimus concentrations were generally maintained within the protocol-defined target range of 3–8 ng/mL. Occasional deviations occurred in a small number of patients but were transient and corrected with dose adjustments.

### Primary Efficacy Outcome

No patient achieved an objective lesion response (predefined as a decrease of at least 30% in diameter) within six months, but two patients showed a regression of the lesion of >10% (patient 01-02: -11.6%, patient 01-05: -15.2%), and 10 patients a regression of between 0% and 10%. The relative changes in max. diameter over the study period are shown in **Figure 1** and **Suppl. Figure 1**. Thus, the predefined radiological endpoint was not met, as 0% of patients (90% CI 0–15.3%, 95% CI 0–18.5%) achieved CR or PR. Consistently, both local investigators’ assessments and the central MRI review confirmed an absence of any radiologic tumor shrinkage meeting RECIST 1.1 response criteria for PR. At the nine-month / post-treatment MRI assessment, none of the patients met the criteria for regression, lesions of 8 patients increased in size, one of which fulfilled the criteria for progressive disease (compared to the nadir of lesion size at the six-months assessment).

**Figure 1.**
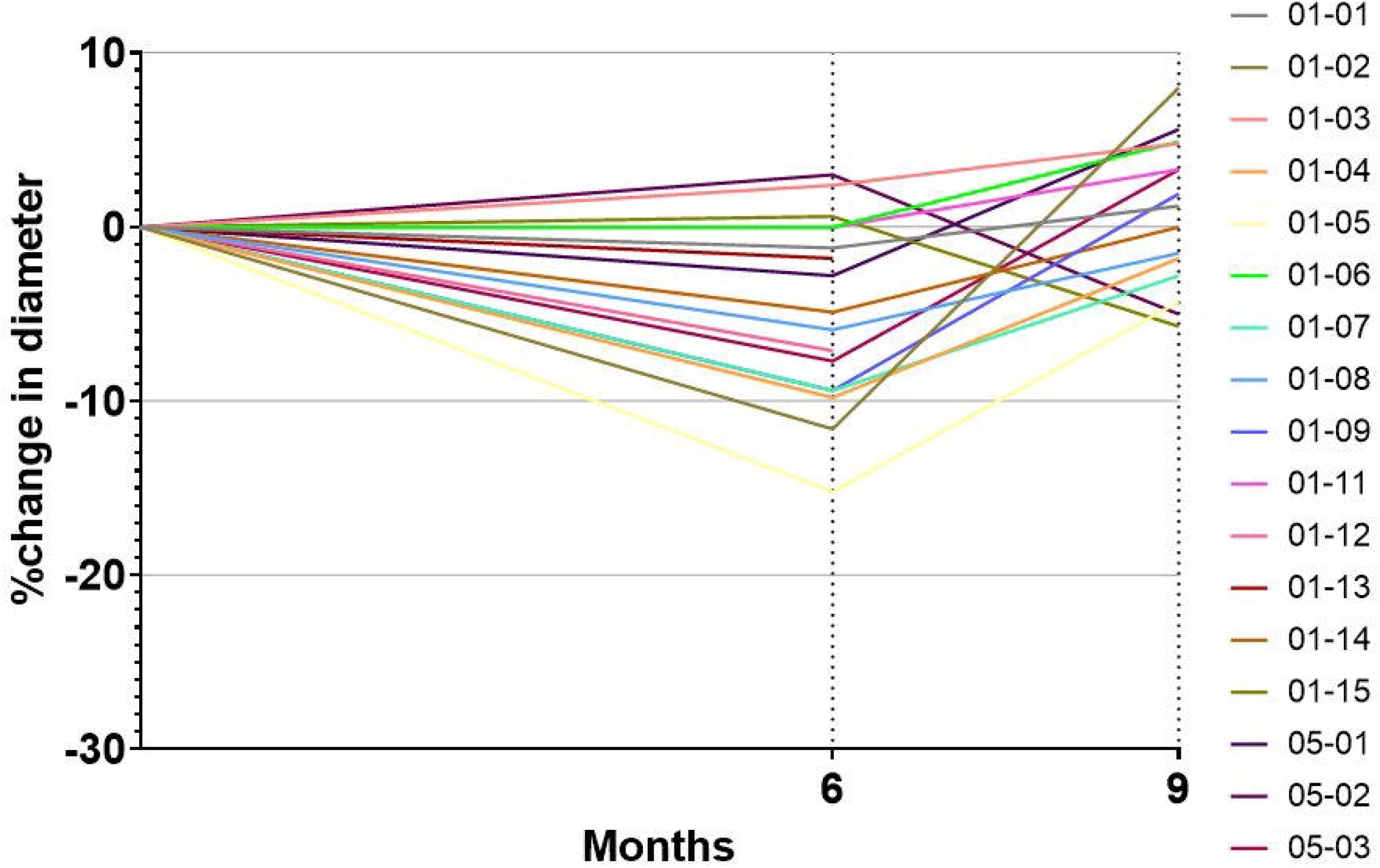
Individual percentage change in lesion size over time assessed by central MRI. Percentage change in lesion size relative to baseline (T0 = 0%) for each patient at 6 and 9 months, based on central MRI assessment. Changes are expressed as relative percentage differences from baseline. In cases of missing follow-up imaging, data are considered not assessed (n.a.) and omitted from the analysis at the respective time point. Due to early study discontinuation before 6 months, patient 01-10 is not represented in the graph.

Despite the lack of radiological response, stable disease was the predominant outcome: by six months, 17 of 18 patients (94%) achieved stable disease as their best response (one patient withdrew from the study before the six-months assessment). Notably, no radiological disease progression was observed during the six-months treatment period, despite 10 of 18 patients having been described as having progressive disease at screening (however, without further information on the dynamics of prior disease progression). No new lesions appeared in any patient while on therapy **(Table 3)**.

**Table 3.** Primary Efficacy Outcome at Month 6 (RECIST 1.1)

| Response Category | n/N | % |
| --- | --- | --- |
| Complete Response (CR) | 0/18 | 0.0 |
| Partial Response (PR) | 0/18 | 0.0 |
| Stable Disease (SD) | 17/18 | 94.4 |
| Progressive Disease (PD) | 0/18 | 0.0 |
| Not evaluable | 1/18 | 5.6 |

Lesion response assessed by magnetic resonance imaging according to RECIST version 1.1. Note: Percentages are based on the full analysis set (N=18).

### Secondary Outcomes

A centralized, blinded photographic assessment was performed to evaluate visible changes over time and record the mean cosmetic response score (0–100 scale, where 0 indicates no visible change and 100 complete resolution). Visible components of the target lesions remained largely unchanged during sirolimus therapy. Overall, only a moderate aesthetic improvement of 8.3 points (SD ±13.9) at six months and 7.4 points (SD ±8.3) at nine months was observed.

Quality-of-life outcomes showed slight improvement in adults but not in pediatric patients. Among adult participants who completed the WHOQOL-BREF questionnaire at month six (n=5), mean difference to baseline at month six was 9.3 (95%-CI: -0.9 to 19.5) in the physical Health domain and 9.2 (95%-CI: 0.7 to 17.7) in the psychological domain.

For pediatric patients, no relevant change in total scores in the KINDL questionnaires was observed over the study period.

Performance status, assessed using the Lansky (pediatric) or Karnofsky (adult) scale, improved modestly. The combined performance score increased by a mean of 3.5 points (95%-CI: 1.0 to 6.1) at six months and 5.7 points (95%-CI: 2.7 to 8.7) at nine months, reflecting a small improvement in daily functioning by study end. **(Table 4)**

**Table 4.**
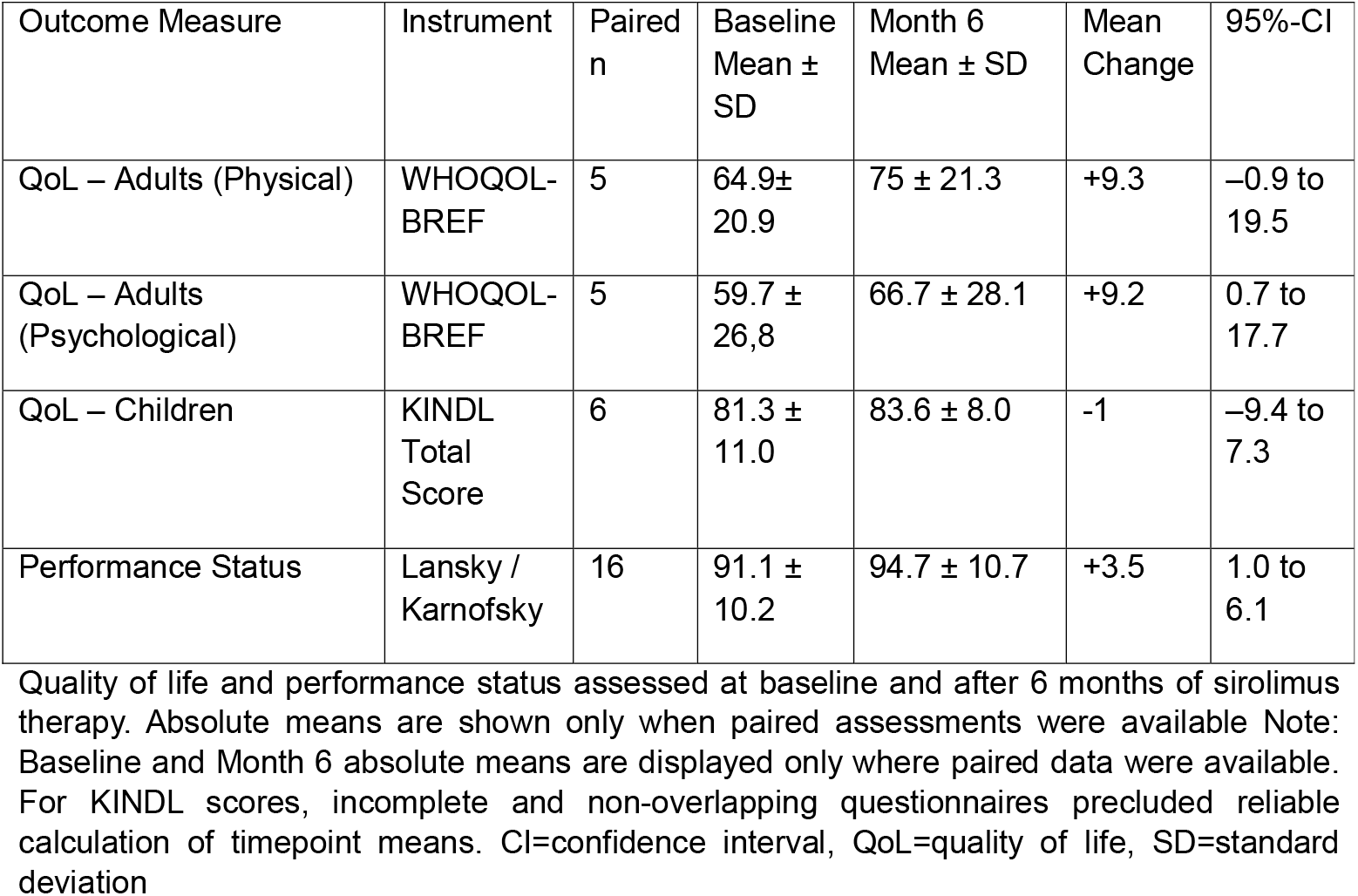
Quality of Life and Performance Status Over Time.

| Outcome Measure | Instrument | Paired n | Baseline Mean ± SD | Month 6 Mean ± SD | Mean Change | 95%-CI |
| --- | --- | --- | --- | --- | --- | --- |
| QoL – Adults (Physical) | WHOQOL-BREF | 5 | 64.9± 20.9 | 75 ± 21.3 | +9.3 | –0.9 to 19.5 |
| QoL – Adults (Psychological) | WHOQOL-BREF | 5 | 59.7 ± 26,8 | 66.7 ± 28.1 | +9.2 | 0.7 to 17.7 |
| QoL – Children | KINDL Total Score | 6 | 81.3 ± 11.0 | 83.6 ± 8.0 | -1 | –9.4 to 7.3 |
| Performance Status | Lansky / Karnofsky | 16 | 91.1 ± 10.2 | 94.7 ± 10.7 | +3.5 | 1.0 to 6.1 |

Quality of life and performance status assessed at baseline and after 6 months of sirolimus therapy. Absolute means are shown only when paired assessments were available Note: Baseline and Month 6 absolute means are displayed only where paired data were available. For KINDL scores, incomplete and non-overlapping questionnaires precluded reliable calculation of timepoint means. CI=confidence interval, QoL=quality of life, SD=standard deviation

Pain levels remained largely unchanged throughout the trial but were already low at screening (mean pain score of 1.4 on a scale of 0 to 10).

Neuropsychological outcomes using the Strengths and Difficulties Questionnaire (SDQ) also showed no relevant change, neither in parent-reported nor in self-reported SDQ total difficulties scores.

Laboratory biomarkers (IGF-1 and IGFBP-3) were within the normal range at baseline and remained normal throughout the study period.

2/18 of the patients showed coagulation abnormalities with lowered fibrinogen, patient 01-01 did not show improved coagulation parameters during therapy, patient 01-02’s D-dimers and fibrinogen levels normalized during the therapy but worsened again after end of therapy.

### Safety

Sirolimus was well tolerated, consistent with its established safety profile. At least one adverse event occurred in all patients, though none were severe. Sirolimus-related adverse events were reported in 10 of 18 patients (55.6%) and included aphthous ulcers (9 patients, 50%), nausea, diarrhea, and acne (2 patients each, 11.1%), and upper abdominal pain (1 patient, 5.6%). Of note, EBV PCR was positive in two patients at screening and normalized during follow-up. One additional patient had a positive EBV PCR at month six; however, no subsequent study visit was available to document normalization. Blood pressure remained normal during the study.

### Follow-up after end of study

After completion of the study protocol, we opted to perform further follow-up of the patients in our registry of vascular anomalies. Six patients restarted sirolimus after a median of 5.7 months (range 1-12 months) after the end of the study treatment with sirolimus, including patient 01-10, who terminated the study after <1 month of therapy. Two additional patients noticed worsening symptoms but deferred restarting of sirolimus. Patient 01-01, who did not benefit from sirolimus treatment, was switched to alpelisib at another center and was then lost to follow-up at our hospital. Six patients were treated with sclerotherapy including bleomycin-electrosclerotherapy with variable efficiency, one patient was treated with repeated embolization. One patient could successfully be weaned from the tracheostoma approx. two years after the study, without restarting sirolimus.

## DISCUSSION

In this open-label, multicenter phase II study, we aimed to evaluate the efficacy and safety of sirolimus in patients with overgrowth with and without vascular malformations. The study showed that sirolimus did not reach the predefined radiological threshold for shrinkage of overgrowth lesions of ≥30% within the six-months treatment period; however, neither did any patient have progressive disease. This finding is consistent with previous studies investigating mTOR inhibition in PROS and related vascular anomalies. In a prospective cohort of 39 patients with PROS, Parker et al. reported modest reductions in tissue volume, with a mean volume reduction of approximately 7% after six months of therapy, despite achieving biochemical target inhibition [2]. Similarly, in the seminal prospective trial by Adams et al. involving complex vascular anomalies, no complete radiological responses were observed, while most patients demonstrated either partial improvement or disease stabilization after six to 12 months of treatment [4]. Consistently, a randomized observational trial in children with slow-flow vascular malformations demonstrated significant improvements in pain, bleeding, oozing, and quality of life, whereas MRI-based lesion volume remained largely unchanged and no complete radiological regressions were reported [3]. More recent multicenter series further support sirolimus as an effective therapeutic option for complicated vascular anomalies, while emphasizing the importance of careful toxicity monitoring and the need for optimized response criteria that better capture clinically meaningful benefits beyond simple lesion size reduction [5,12]. The European VASE trial investigating the use of sirolimus in vascular anomalies reports a clinical response rate of >80% but changes in lesion size are not reported [13]. Together, these studies indicate that meaningful lesion shrinkage under sirolimus is uncommon, but patients may experience symptomatic benefit and disease stabilization. Stable disease during treatment may still represent a clinically relevant signal in patients with previously progressive overgrowth. However, pretreatment growth kinetics were not prospectively or systematically captured in this trial, representing an important limitation. This consideration may be particularly relevant in younger children, in whom preventing additional disproportionate growth during development could be clinically meaningful even in the absence of RECIST-defined shrinkage.

The relatively short treatment duration of six months in our study may further explain the absence of radiological regression. Segmental overgrowth lesions typically evolve over years, and reversal of established tissue architecture may require pathway suppression for a prolonged period. In longer-term observational cohorts, continued sirolimus therapy beyond six months has been associated with sustained clinical benefit in patients with complicated vascular anomalies, although without dramatic volumetric regression [4]. Therefore, a six-months study period may be insufficient to capture the full therapeutic potential of mTOR inhibition, particularly when radiological endpoints are used as the primary efficacy measure. RECIST 1.1 was chosen as the primary outcome to utilize a standardized and widely recognized tool to standardize evaluation of response. Nonetheless, its applicability to complex vascular and overgrowth lesions seemed to be limited. These lesions often are infiltrative, poorly circumscribed and composed of vessels, adipose tissue and fibrous stroma and may show complex patterns of response. Additionally, lesions might rather exhibit a decrease in thickness instead of the length of the lesion, which might not be reflected in measuring the longest diameter. Thus, even clinically meaningful changes may not translate into a ≥30% decrease in the longest diameter and overgrowth of multiple tissue types may respond differently to sirolimus treatment. Recent studies have highlighted the poor sensitivity of linear measurements for detecting changes in slow-flow vascular malformations and PROS lesions, suggesting that volumetric analysis or disease-specific scoring systems may be more appropriate [3,5]. However, even with a more exact volumetric assessment, the predefined radiological readout may be missed despite significant clinical benefit, as was the case with the EPIK-P2 trial on alpelisib in PROS patients [14].

Despite the lack of objective radiological responses, sirolimus treatment was associated with a favorable clinical response. We could observe modest improvements in Lansky/Karnofsky performance status both in pediatric and adult patients, the latter also demonstrated improved psychological QoL scores. These findings align with previous reports emphasizing symptomatic and functional improvement as key benefits of sirolimus therapy and underlining the need for specialized outcome scales such as the OVAMA questionnaire [15]. Engel et al. demonstrated that over 90% of patients with PROS-associated vascular malformations experienced improvement in at least one clinical symptom, including pain, bleeding, or functional impairment, despite minimal radiological change [16]. Notably, resolution of coagulopathy and bleeding complications occurred even in the absence of lesion shrinkage [16], highlighting the dissociation between radiological and clinical outcomes.

Importantly, several patients in our cohort expressed a desire to resume or continue sirolimus therapy after study completion, further suggesting a perceived subjective benefit that may not be fully captured by imaging-based endpoints. This observation underscores a critical limitation of relying solely on radiological response criteria in patients with complex overgrowth, as a treatment response may rather manifest as pain control, functional improvement, relief of aesthetic burden, and increased health-related quality of life.

Future studies should incorporate comparative designs whenever feasible, e.g. utilizing randomized delayed-start designs. Additionally, future clinical trials should consider focusing on patient-centred outcome measures alongside radiological assessments to more accurately evaluate treatment efficacy.

Sirolimus demonstrated a favorable safety profile in both pediatric and adult patients, with no new or unexpected safety signals. All adverse events observed were consistent with previously reported side-effects of mTOR inhibitors. Most events were mild to moderate in severity and manageable with supportive care or dose adjustments. No treatment-related deaths, serious adverse events, or irreversible toxicities were reported. These findings generally are in line with previously published data. In the prospective trial by Adams et al., a small proportion of patients discontinued sirolimus due to adverse events, and no treatment-related mortality was observed. [4] Engel at al. reported transient cytopenias and infections as the most common adverse effects, with no life-threatening complications [16]. However, it is noteworthy that Parker et al. reported grade ≥3 toxicities in approximately one-third of PROS patients, emphasizing the importance of careful patient selection and monitoring [2]. Overall, the benefit–risk balance of sirolimus appears favourable in patients with severe, progressive overgrowth syndromes who have limited therapeutic alternatives, provided that appropriate surveillance is ensured.

This study has several limitations: First, the study included a small, heterogeneous patient cohort, reflecting the rarity and phenotypic diversity of overgrowth diseases with or without a vascular anomaly. Second, the absence of a control arm precludes definitive attribution of disease stabilization to sirolimus alone, as natural disease variability cannot be excluded. Third, the 6-month treatment duration and follow-up were relatively short. Overgrowth syndromes represent chronic conditions, and longer observation periods are needed to assess sustained efficacy, late toxicity, and disease rebound after treatment cessation. Fourth, molecular confirmation was unavailable in 8/18 participants. Although eligibility in this study was based on clinical and radiological diagnosis and the protocol explicitly allowed inclusion independently of genetic background, incomplete genotyping limits biological precision, precludes exploratory genotype–response analyses, and complicates comparison with genotype-directed approaches. Finally, radiological response according to RECIST 1.1 may not fully capture clinically relevant changes in non-neoplastic lesions, limiting the sensitivity of the primary endpoint.

We propose future research may explore the following avenues. A longer treatment duration and earlier therapeutic intervention may enhance disease control and prevent irreversible tissue changes. The emergence of PI3Kα inhibitors provides an upstream targeted option that may achieve more robust volumetric responses in genetically confirmed PROS. In the EPIK-P1 study, 27% of patients had a radiologic response at 24 weeks, many with responses lasting at least 12 months. Further, the majority reported symptom improvement irrespective of radiologic response – this disconnect of radiological and clinical response was also observed in the EPIK-P2 trial [14] and occurred similarly in our cohort. Additional case series of alpelisib treatment in children and infants with PROS described marked reduction of overgrowth and functional gains, including in patients previously treated with sirolimus [17– 19]. Future trials of genotype-directed agents should incorporate prospective molecular confirmation wherever feasible to enable biologically more homogeneous cohorts and more informative treatment comparisons. Combination therapies targeting multiple nodes of the PI3K/AKT/mTOR pathway may represent a promising strategy; however, further preclinical evidence is needed prior to exploring this strategy in clinical trials.

Finally, the development and implementation of standardized, disease-specific outcome measures that incorporate functional, symptomatic, and patient-reported outcome endpoints as well as radiological assessment will be essential to advance therapeutic response evaluation in overgrowth with or without a vascular anomaly.

## CONCLUSION

Sirolimus is a safe and well-tolerated therapeutic option for patients with overgrowth with or without vascular anomalies. Although no participant achieved RECIST-defined lesion reduction over the 6-months treatment period, disease remained stable in most evaluable participants, and modest improvements were reported in selected patient-reported and functional outcomes. The fact that a relevant number of patients elected to resume sirolimus after study completion suggests that clinical benefit was not fully captured by imaging alone. Given its favorable safety profile, comparably low cost, and established long-term use, sirolimus remains a valuable treatment option for patients with overgrowth with and without vascular anomalies. Future studies should address optimal treatment duration, timing of intervention, and the potential for comparative trials or combination strategies with targeted agents such as PI3Kα inhibitors to optimize long-term outcomes in this complex patient population.

## Supporting information

Waterfall plots of change in lesion diameter at 6 months and at 9 months (3 months after discontinuation of sirolimus treatment).

## Data Availability

The datasets generated and/or analyzed during the current study are not publicly available due to privacy and ethical restrictions but may be available from the corresponding author upon reasonable request.

## ACKNOWLEDGEMENTS

The study was funded by the German Research Foundation (DFG RO 1263/10-1), after a seed funding in the Clinical Trials Program of the Medical Faculty of the University Freiburg. The study drug was provided by Pfizer Germany through an independent research grant. We acknowledge the Center for Vascular Anomalies at the Freiburg Center for Rare Diseases and the Hilda Biobank at the Department of Pediatrics and Adolescent Medicine, Freiburg, Germany for their contribution. Both participating centers are members of the German Reference Network for Vascular Anomalies. FGK was supported by the charity cycling tour “Tour der Hoffnung” and received funding from the German Federal Ministry of Education and Research (BMFTR): EJPRD Joint Transnational Call 2023, NARRATIVE; FKZ 01GM2405). We thank Maria Huber for help in planning the clinical trial.

## AUTHOR CONTRIBUTIONS

Gabriella Cericola: visualization, writing – original draft preparation, writing – review and editing

Beate Häberle: investigation, writing – review and editing

Julia Haehl: investigation, writing – review and editing

Sebastian Berg: central radiological assessment, writing – review and editing

Simon Leschka: central radiological assessment, writing – review and editing

Susanne Grüninger: conceptualization, project administration

Claudia Schmoor: statistical analysis Sebastian Wirtz: statistical analysis

Annegret Holm: prescreening, writing – review and editing Charlotte M. Niemeyer: resources

Alexander Puzik: investigation, writing – review and editing

Martin Zenker: genetic analyses, writing – review and editing

Moritz Wildgruber: investigation, writing – review and editing

Jochen Rößler: conceptualization of the study, funding acquisition, writing – original draft preparation

Friedrich G. Kapp: conceptualization of the study, investigation, funding acquisition, visualization, writing – original draft preparation, writing – review and editing, supervision, resources.

All authors provided intellectual input, critical feedback, discussed results, and approved the final manuscript.

## SUPPLEMENTARY MATERIAL

**Suppl. Figure 1: Waterfall plots of change in lesion diameter**

