## Supplementary figures and images for "Sirolimus in Patients with Overgrowth, with or without Vascular Malformations: The SIPA-SOS Study"

### Waterfall plots of change in lesion diameter at 6 months and at 9 months (3 months after discontinuation of sirolimus treatment).

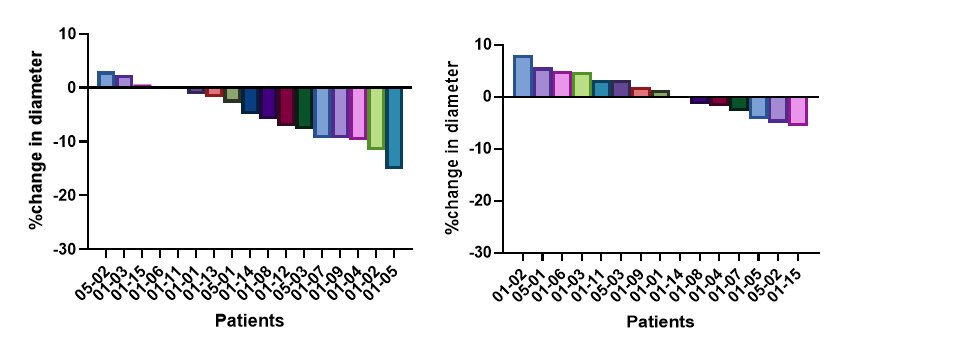
